# Understanding the impact of incontinence on Veterans’ self-management strategies, quality of life and treatment experiences

**DOI:** 10.1101/2020.11.25.20231506

**Authors:** Linda Cowan, Sarah Bradley, Andrew R. Devendorf, Lelia Barks, Tatiana Orozco, Angelina Klanchar, Jennifer Hale-Gallardo

## Abstract

**Background:** Urinary and fecal incontinence contribute to significant quality of life impairments for patients and caregivers. Preliminary research suggests that incontinence rates may be higher among Veterans. However, few studies have examined incontinence experiences among Veterans and their caregivers.

**Objectives:** We aimed to demonstrate the feasibility of conducting a one-year, telephone survey with Veterans and their caregivers to help inform larger studies. By including both Veteran and caregiver perspectives, we explored how incontinence impacts Veterans’ self-management strategies, quality of life, and treatment experiences.

**Design:** We used a mixed methods design, employing quantitative (i.e., cross-sectional survey) and qualitative approaches (i.e., semi-structured interviews).

**Participants:** Our sample included 64 Veterans with urinary incontinence, fecal incontinence, or mixed incontinence, and 36 caregivers. A subset of 18 Veterans and 8 caregivers completed semi-structured interviews.

**Methods:** Data were collected via telephone surveys over the course of 1-year from a small research team at the Veterans Health Administration (VHA). Participants completed measures about the Veteran’s incontinence severity, quality of life, and VHA treatment experiences. Interviews asked participants about their perceptions and satisfaction in receiving treatment for the Veterans’ incontinence. Qualitative themes were extracted using a Rapid Assessment Process model.

**Results:** Veterans’ self-reported physical quality of life correlated negatively and significantly with both urinary and fecal incontinence severity, as well as negatively and significantly with urinary continence bother (rs range: −.36 to −.47, ps < .01). Veterans’ mental quality of life correlated negatively and significantly with urinary incontinence bother (r = −.43, p < .001). About 67% Veterans experienced incontinence symptoms for 5-years or more, yet 44% waited at least 1-year to discuss incontinence with a VHA provider. Most Veterans (92%) reported speaking with a VHA provider about incontinence, while only 42% reported speaking with a non-VHA provider. Qualitative findings revealed that, upon speaking to a provider, Veterans felt comfortable but also desired more incontinence education from their providers, including obtaining more appropriate and tailored treatment options. While most Veterans followed their provider’s treatment recommendations, some felt unsatisfied with treatments that they perceived as embarrassing.

**Conclusions:** Recruiting a substantial sample of Veterans with incontinence, and their caregivers, is feasible using our recruitment methods, which can inform larger studies. Our study revealed that incontinence symptoms interfere significantly with the quality of life of Veterans and their caregivers. Intervening at the healthcare level by educating providers and systematizing inquiry into incontinence for higher risk populations would be fruitful to explore.

**Contribution of the Paper:** *What is already known about this topic?:*

- Urinary incontinence is more prevalent than fecal incontinence, and both are associated with significant impairments in physical, mental, and social functioning.
- Urinary and fecal incontinence become more prevalent with older age and additional chronic health conditions.
- There is a lack of incontinence diagnoses documented in Veteran’s medical records, even when incontinence is present. Few studies have reported on caregiver burden related to incontinence care.

*What this paper adds:*

- This study is the first to report on the potential delay between Veteran’s experiencing symptoms of incontinence and informing their healthcare provider or caregiver about those symptoms (sometimes 5-years or more).
- Insights on Veteran and caregiver satisfaction with incontinence care can guide healthcare interventions to improve incontinence care.
- Both Veteran quality of life and caregiver burden correlated significantly and negatively with satisfaction with incontinence treatments tried and number of treatments tried. Only 21% of Veterans were satisfied with the treatment plan they were given, suggesting a potential knowledge gap or opportunity for improvement in healthcare provider approaches to incontinence management.

---

Over 15 million American adults experience urinary incontinence (UI), fecal incontinence (FI), or both (Markland et al., 2010). Both UI and FI are associated with significant impairments in physical, mental, and social functioning (Chang, Gonzalez, Lau, & Sier, 2008; Mardon, Halim, Pawlson, & Haffer, 2006). However, few studies have investigated incontinence among U.S. Veterans, a population in which incontinence is likely underreported and undertreated (Smoger et al., 2000). In a literature review from 1990-2019, there were only 9 research articles published which reported on incontinence among Veterans. Most of these studies explored UI and FI separately, rather than together. Consequently, healthcare providers lack an understanding about the experiences of incontinence among Veterans and their caregivers. Knowledge about Veterans’ perceptions and management of incontinence would assist healthcare providers with the screening and treatment of incontinence. To fill this gap, we report on findings from a 2019 pilot study that employed telephone surveys and interviews to investigate the experiences of both Veterans with incontinence and their caregivers. By including both Veteran and caregiver perspectives, we explore how UI and FI impacts Veterans’ self-management strategies, quality of life, and treatment experiences.

## Epidemiology of Incontinence

The prevalence rates of UI and FI vary by definitions of incontinence, study population, and research designs (Hampel et al., 1997; Ditah et al., 2014). About 10% to 20% of all women, and an estimated 20% of all men, experience UI in the U.S. (Lukacz et al., 2017; Markland et al., 2010). The prevalence of FI spans 8.3%-20% among community-dwelling, ambulatory adults (Johanson & Lafferty, 1996; Whitehead et al., 2009). UI and FI become more prevalent with older age and additional chronic health conditions (Ditah et al., 2014; Hampel et al. 1997; Rabinowitz, Martin, Montague, & Hirdes, 2011; Martin, Rabinowitz, & Montague, 2012; Komesu et al., 2016). Urinary incontinence is most commonly associated with four urologic conditions: benign prostatic hypertrophy; lower urinary tract dysfunction; kidney stones; and urinary tract infections. Fecal incontinence is more prevalent in individuals with inflammatory bowel disease, celiac disease, diabetes, and irritable bowel syndrome (Madoff et al., 2004; Menees, Almario, Spiegel, & Chey, 2018).

Existing prevalence estimates of incontinence among Veterans are inconsistent. In three Veterans Health Administration (VHA) outpatient primary care clinics, 32.3% of male Veterans reported UI symptoms at least once over the previous 12 months, and 13.8% reported UI symptoms on a weekly basis (Smoger,’ Felice, & Kloecker, 2000). Vaughan et al. (2014) found that, among men aged 55-years or younger, those with military exposure had 3 times greater odds of UI compared to men without military exposure. In female Veterans, UI prevalence increases with age and comorbidities like PTSD, depression, and sexual trauma (Klausner et al., 2009; Bradley et al., 2017; Bradley et al., 2012). While these studies indicate that UI is more common in Veterans, Anger et al. (2008) found a lower *documented* UI prevalence among Veterans in outpatient clinics, with just 2.2% of female Veterans and only 0.5% of male Veterans having a UI diagnosis documented in their medical record.

Current data on Veterans’ FI prevalence nationwide is sparse, but a recent article indicated 31% of Veterans in a primary care clinic reported accidental bowel movements within the previous 30 days compared to 41% of Veterans referred to a gastroenterology (GI) clinic (Hosmer, Saini & Menees, 2019), with an average age of all participants of 59-years-old. These findings demonstrate the need for further investigation into incontinence risk factors that are specific to military service members, including younger and older Veterans (Vaughan et al., 2014).

## Impact of Incontinence and Treatment Barriers

Both UI and FI are detrimental to an individual’s physical, psychosocial, and financial well-being (Hawkins et al., 2011; Markland et al., 2011). Not only does incontinence increase the risk for rashes, skin breakdown, and pressure ulcer formation (Beeckman, Van Lancker, Van Hecke, & Verhaeghe, 2014), many patients also experience shame, embarrassment, and a lack of confidence (Garcia, Crocker, & Wyman, 2005; Devendorf et al., 2020). Many individuals with UI or FI may avoid disclosing their incontinence to their caregiver, or delay seeking incontinence treatment. Treatment barriers include feelings like embarrassment, lack of knowledge about treatments, and beliefs that incontinence is “normal” after events like a stroke (Taylor & Cahil, 2018; Devendorf et al., 2020). Additionally, healthcare providers may not query their patients about symptoms (Strickland, 2014; Hosmer, Saini & Menees, 2019).

Few studies have investigated factors related to Veterans’ perceived symptom burden (e.g., severity), treatment-seeking behaviors (e.g., time to initiate treatment), and healthcare experiences regarding UI or FI (e.g., satisfaction with care). Relatedly, there are no incontinence studies, to our knowledge, that include caregivers’ reports or experiences in relation to Veterans’ incontinence. Caregivers may provide additional, and potentially more forthcoming, accounts of Veterans’ experiences with a topic that is often perceived as embarrassing (Taylor et al., 2013). It is also important to understand the impact of incontinence on the caregivers’ quality of life. These gaps must be addressed before developing research and educational interventions that could improve the practice and quality of incontinence care for Veterans within the healthcare system.

## The Current Study

Because little research exists on incontinence among Veterans, we conducted a pilot study in preparation for a larger, national survey to address these research gaps. Our pilot study had three goals. First, we aimed to evaluate the telephone survey and interview questions, the effectiveness of the recruitment methods, and the resources needed for a larger future study. To accomplish this goal, the study aimed to recruit 100 total subjects. Second, we aimed to describe the demographic and clinical characteristics of Veterans with incontinence, especially regarding their symptom severity, treatment-seeking behaviors, healthcare experiences, and incontinence management. Third, we aimed to use both quantitative and qualitative data to gain insight into Veterans’ experiences with incontinence treatment (e.g., satisfaction with care), and the experiences and perceptions of their caregivers at home.

## Methods

### Design

This study used a mixed methods design, employing both quantitative and qualitative approaches. In part 1 of the study, we administered a telephone survey to Veterans with incontinence and/or their caregivers (defined as an individual who assists with incontinence care and lives with the patient). These surveys consisted of multiple choice, Likert scale responses, and several open-ended text questions. In part 2, a subset of our overall sample (18 Veterans and 8 caregivers) participated in semi-structured qualitative interviews, which utilized open-ended probing questions in an interview guide format over the phone. These qualitative interviews allowed us to gain additional in-depth information about the lived experience with incontinence.

### Recruitment and Procedure

The target sample size was approximately 100 Veterans and/or their caregivers. This sample was determined due to feasibility reasons, as the study had a one-year duration (January 2019 to December 2019). Following IRB approval from the “DEIDENTIFIED FOR REVIEW Board, (Project #00038498)” participants were recruited. Figure 1 in the Supplementary Material provides information about our recruitment.

**Figure 1.**
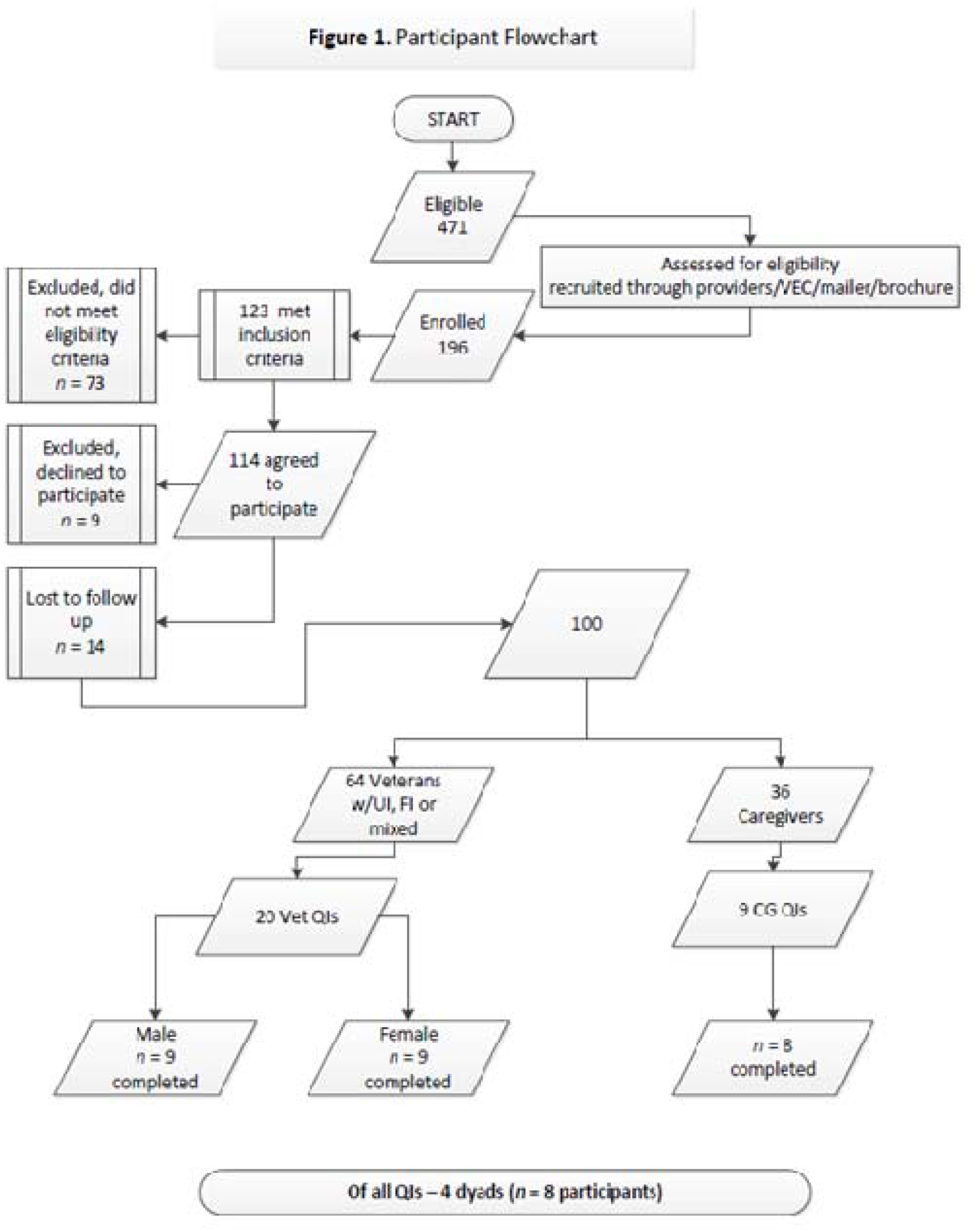
Participant Flowchart.

Altogether, 471 community-dwelling Veterans were approached for recruitment through several methods, including through Veterans’ providers (who were emailed by the research team), a brochure and presentation to the DEIDENTIFIED FOR REVIEW Veteran Engagement Group, snowball referrals, an invitation letter and study brochure mailout, and brochures left in primary care clinics. Interested Veterans and/or their caregivers contacted the research team by phone or return mail, provided verbal informed consent, were screened for eligibility, and then were scheduled for a phone survey with a trained nurse. A subset of participants was given the opportunity to participate in the qualitative interviews.

### Participants

The following inclusion criteria were applied: male and female Veterans aged 18-100 years old with UI or FI, who were able and willing to participate in a telephone survey, residing in either single family home or Medical Foster Home, and their primary, residential, direct caregivers. The following exclusion criteria were applied: Veterans or their caregivers who did not agree to answer phone survey questions, did not speak English, or those with any disability that prevented them from communicating over the phone or giving their own verbal consent. These exclusions of communication did not prevent their caregivers from participating in the caregiver portion of the survey if they wished. Individuals enrolled in another research study at the same time and Veterans residing in institutional environments (e.g., nursing homes or assisted living facilities) were also excluded. Of the 471 Veterans approached for recruitment, 275 contacted did not respond, 73 did not meet inclusion criteria, 9 declined to participate, and 14 were lost to follow-up.

Our final sample includes 64 Veterans with UI, FI, or mixed incontinence, and 36 willing caregivers, including 13 Veteran-caregiver dyads. A subset of these Veterans (n=20) and caregivers (n=9) agreed to participate in the part qualitative interviews; of these, one Veteran-caregiver dyad ultimately declined, and one Veteran was unintelligible by phone, leaving 26 participants (18 Veterans; 8 caregivers) who completed the qualitative portion.

### Quantitative Measures

#### Michigan Incontinence Symptom Index (MISI)

The MISI assessed the severity of participants’ UI. The MISI consists of 10-items using Likert scale responses that can be constructed as two domains: Total MISI Domain (consisting of subdomains for stress urinary incontinence, urgency urinary incontinence, and pad use) and a Bother Domain. The MISI has demonstrated strong psychometric properties among patients with UI (Suskind et al., 2014).

#### Adapted Fecal Incontinence Severity Index (FISI)

The FISI assessed the severity of participants’ FI. The FISI normally consists of 4 items that query participants on the frequency of incontinence for gas, mucus, liquid stool, and solid stool. The total score sums all items and varies from 0 to 61, where higher scores indicate higher perceived severity of the fecal incontinence (Rockwood et al., 1999). However, due to our research interests, we queried participants only on the liquid stool and solid stool items (leaving out questions related to passing gas or mucous), leaving a total score range of 0-37.

#### Rand 12-Item Health Survey (VR-12)

The VR-12 was only administered to Veterans and measured their health-related quality of life. The wording on some of the VR-12 items was edited to refer specifically to their incontinence symptoms (rather than symptoms more broadly, which could be in reference to other conditions). The VR-12 includes 12-items on a Likert scale, and it provides a physical and mental health component using standardized scores. Specifically, T-scores were computed using a SAS scoring algorithm provided by Spiro et al. (2004), and then updated to reflect more current population norms (Selim et al., 2009). VR-12 is norm-scored such that the general U.S. population scores have a mean of 50 (*SD*=10). The VR-12 has demonstrated strong psychometric properties (Kazis et al., 2006; Kazis et al., 2006).

#### Zarit [Caregiver] Burden Interview-12 (ZBI)

The ZBI was only administered to caregivers and assessed perceptions of the burden of caring for incontinence. The ZBI includes 12 questions on a 0-4 Likert scale. Total scores range from 0-48, with higher scores indicating worse perceived burden. Research has recommended that scores of 10-19 indicate mild to moderate burden, while scores of 20 or more indicate severe burden. The ZBI is commonly used in population-based studies and has demonstrated strong psychometric properties (Bedard et al., 2001; Higginson et al., 2010).

#### Additional Incontinence Measures

Participants were queried about Veterans’ duration of incontinence symptoms, incontinence management, treatment experiences (e.g., treatments recommended, treatments tried) in VHA health care settings, duration in symptom onset and treatment initiation, and satisfaction with VHA care.

### Qualitative Procedure and Methods

Interview questions were prepared and piloted by a qualitative expert with items suggested by the literature (see Appendices A-D for the qualitative interview guides and specific questions). After taking the telephone survey, participants who agreed to complete the qualitative interview were contacted later and interviewed by one of our team’s qualitative investigators. Interview questions concerned participants’ knowledge of incontinence, their experiences seeking treatment for incontinence, interactions with providers, symptom management, and satisfaction with incontinence products. Interviews were recorded, transcribed verbatim, and checked for accuracy.

### Data Analytic Plan

#### Quantitative Analysis

Data were stored and analyzed with VA REDCap, Microsoft Excel, SAS, and R/RStudio. Descriptive statistics and zero-order correlations (employing listwise deletion for the few missing datapoints) were conducted to examine relationships amongst variables. We provide descriptive information about Veterans’ perceived severity, duration, management, and treatment utilization in relation to incontinence. We also provide correlations to describe relationships amongst these variables. An alpha value of .05 denoted statistical significance.

#### Qualitative Analysis Plan

For this manuscript, our qualitative analysis focused on understanding Veterans’ and their caregivers’ perceptions and satisfaction in receiving treatment for the Veterans’ incontinence. We have provided detailed documentation of the qualitative methods in a prior investigation, which focused on the topics of stigma, incontinence, and Veterans experiences (see DEIDENTIFIED FOR REVIEW, in press). In this work, we provide a more complete description for our quantitative methods and only a cursory description of our qualitative methods.

All interviews were transcribed verbatim and checked for accuracy. Responses were entered into a rapid-assessment matrix. The Rapid Assessment Process model is a systematic and iterative analytical approach to efficiently identify emergent themes from qualitative data (Beebe, 2001). The analysis included the following steps: (1) creation of domain names that correspond with interview questions; (2) development of a note-taking template using interview guide and domain names; (3) conduction of debriefings and refinement of notes following interviews and observations; (4) categorization of responses under corresponding domain names; transfer of notes to an Excel® matrix; (6) review of categorizations and establishment of consensus among the team; and (7) analyzation and summarization of domains for key themes, variations, and information gaps.

## Results

### Quantitative

#### Results Demographic Characteristics

Table 1 provides demographics for Veterans and their caregivers. All Veteran participants experienced UI, with 31 (48.44%) reporting UI only, and 33 (51.56%) experiencing both UI and FI. No Veterans reported experiencing only fecal incontinence. Veterans (n=64) were generally male (65.63%), Caucasian (64.06%), and older (M=65.42; SD=13.65). More than one-third of Veterans (n=27, 42.19%) reported that incontinence ever caused a rash or skin irritation (Table 2).

**Table 1.** Veteran and Caregiver Demographics.

|  | Veteran<br>(n=64) |  | Caregiver<br>(n=36) |  | Total<br>(n=100) |  |
| --- | --- | --- | --- | --- | --- | --- |
|  | *N* | *Mean (SD) or %* | *N* | *Mean (SD) or %* | *N* | *Mean (SD) or %* |
| Age, in years | 64 | 65.42 (13.65) | 35 | 62.74 (13.03) | 99 | 64.47 (13.34) |
| Gender |  |  |  |  |  |  |
| Female | 22 | 34.38% | 33 | 91.67% | 55 | 55.00% |
| Male | 42 | 65.63% | 3 | 8.33% | 45 | 45.00% |
| Ethnicity/Race* |  |  |  |  |  |  |
| African or African-American | 15 | 23.44% | 5 | 13.89% | 20 | 20.00% |
| Asian | 1 | 1.56% | 2 | 5.56% | 3 | 3.00% |
| Caucasian | 41 | 64.06% | 28 | 77.78% | 69 | 69.00% |
| Hispanic or Latino | 6 | 9.38% | 5 | 13.89% | 11 | 11.00% |
| Native American or Pacific Islander | 3 | 4.69% | 3 | 8.33% | 6 | 6.00% |
| Other | 2 | 3.13% | 1 | 2.78% | 3 | 3.00% |

\* Percentages within this variable may add up to >100 due to overlap/multiple responses permitted

**Table 2.** Characteristics of Veterans with Incontinence.

|  | Total (N=64) |  | Urinary (N=31) |  | Both (N=33) |  |
| --- | --- | --- | --- | --- | --- | --- |
|  | *N* | *%* | *N* | *%* | *N* | *%* |
| **Incontinence Duration** |  |  |  |  |  |  |
| <1 month | 0 | 0.00% | 0 | 0.00% | 0 | 0.00% |
| 1 month – 6 months | 1 | 1.56% | 0 | 0.00% | 1 | 3.03% |
| 6 months – 1 year | 1 | 1.56% | 1 | 3.23% | 0 | 0.00% |
| 1 year – 5 years | 18 | 28.13% | 9 | 29.03% | 9 | 27.27% |
| 5+ years | 43 | 67.19% | 20 | 64.52% | 23 | 69.70% |
| Missing | 1 | 1.56% | 1 | 3.23% | 0 | 0.00% |
| **Caused Skin Irritation?** |  |  |  |  |  |  |
| No | 37 | 57.82% | 19 | 61.29% | 18 | 54.55% |
| Yes | 27 | 42.19% | 12 | 38.71% | 15 | 45.45% |
| **Used these to manage skin irritation?**<br>**(Multiple Responses Allowed)** |  |  |  |  |  |  |
| Not Applicable | 39 | 60.94% | 22 | 70.97% | 17 | 51.52% |
| Prescription Meds (Someone Else's) | 0 | 0.00% | 0 | 0.00% | 0 | 0.00% |
| OTC Meds | 8 | 12.5% | 4 | 12.90% | 4 | 12.12% |
| OTC Incontinence Products (e.g., pads or disposable briefs) | 13 | 20.31% | 5 | 16.13% | 8 | 24.24% |
| Protective Creams or Ointments | 18 | 28.13% | 5 | 16.13% | 13 | 39.39% |
| Powders | 3 | 4.69% | 1 | 3.23% | 2 | 6.06% |

Caregivers (n=36) were generally female (91.67%), Caucasian (77.78%), and older (M=62.74; SD=13.03). Most caregivers reported being the spouse of their Veteran (56% of caregivers) and were not currently employed (69% of caregivers). Caregivers varied in the amount of time they spent performing daily personal care related to incontinence, with 10 (27.78%) caregivers reporting 1 hour or less, 16 (44.44%) reporting 1.5 to 4 hours, and 8 (22.22%) reporting 5 to 12 hours per day.

### Clinical Characteristics

#### Veteran Treatment Experiences

Table 3 provides information about Veteran’s treatment seeking behaviors. Most Veterans (92%) had at some point told a VA provider about their incontinence, and 41% told a non-VA provider. While 28% of Veterans experienced incontinence symptoms for 1-5 years, and 67% of Veterans experienced symptoms for 5-years or more, 44% of all Veteran participants waited more than 1-year after the onset of incontinence symptoms to discuss incontinence with their health care provider.

**Table 3.** Frequencies for Items Relating to Healthcare Provider Discussions.

|  | **Total (N=64)** |  | **Urinary (N=31)** |  | **Both (N=33)** |  |
| --- | --- | --- | --- | --- | --- | --- |
|  | *N* | *%* | *N* | *%* | *N* | *%* |
| **Told VA Provider?** |  |  |  |  |  |  |
| *No* | 5 | 7.81% | 1 | 3.23% | 4 | 12.12% |
| *Yes* | 59 | 92.19% | 30 | 96.77% | 29 | 87.88% |
| **Time waited to tell VA provider** |  |  |  |  |  |  |
| Not Applicable | 4 | 6.25% | 1 | 3.23% | 3 | 9.09% |
| < 1 month | 20 | 31.25% | 10 | 32.26% | 10 | 30.30% |
| 1-6 months | 8 | 12.50% | 2 | 6.45% | 6 | 18.18% |
| 6 months – 1 year | 4 | 6.25% | 3 | 9.68% | 1 | 3.03% |
| 1-5 years | 17 | 26.56% | 10 | 32.26% | 7 | 21.21% |
| >5 years | 11 | 17.19% | 5 | 16.13% | 6 | 18.18% |
| **Told Non-VA Provider?** |  |  |  |  |  |  |
| *No* | 38 | 59.38% | 20 | 64.52% | 18 | 54.55% |
| *Yes* | 26 | 40.63% | 11 | 35.48% | 15 | 45.45% |
| **Time waited to tell non-VA provider** |  |  |  |  |  |  |
| Not Applicable | 38 | 59.38% | 20 | 64.52% | 18 | 54.55% |
| < 1 month | 12 | 18.75% | 7 | 22.58% | 5 | 15.15% |
| 1-6 months | 6 | 9.37% | 0 | 0.00% | 6 | 18.18% |
| 6 months – 1 year | 2 | 3.13% | 2 | 6.45% | 0 | 0.00% |
| 1-5 years | 2 | 3.13% | 1 | 3.23% | 1 | 3.03% |
| >5 years | 4 | 6.25% | 1 | 3.23% | 3 | 9.09% |

Tables 1 and 2 in the Supplementary Material provides information about incontinence treatments which were commonly recommended from healthcare providers and those which were tried by Veterans. Veterans reported being recommended (*M* = 3.09, *SD* = 1.72) and trying (*M* = 2.41, *SD* = 1.27) several treatments from healthcare providers. Incontinent products (such as briefs) supplied by the VA (85.94%), medications (64.06%), and physical therapy or specialist therapy (40.63%) were the most recommended treatments from healthcare providers.

Accordingly, incontinent products supplied by the VA (85.94%), medications (60.94%), and physical therapy or specialist therapy (34.38%) were also the most tried treatments by Veterans.

#### Veteran Quality of Life

Table 4 provides descriptive information and zero-order correlations related to Veterans’ symptoms. On the VR-12, Veterans reported having considerable quality of life impairments. Specifically, Veterans had a mean physical component summary T-score of 41.49 (*SD*=9.70), and a mean mental component summary T-score of 42.39 (*SD*=12.27), which is considerably lower than the U.S. population norms (M=50; S*D*=10). Thus, it appears the Veterans in this sample experienced a reduced quality of life.

**Table 4.**
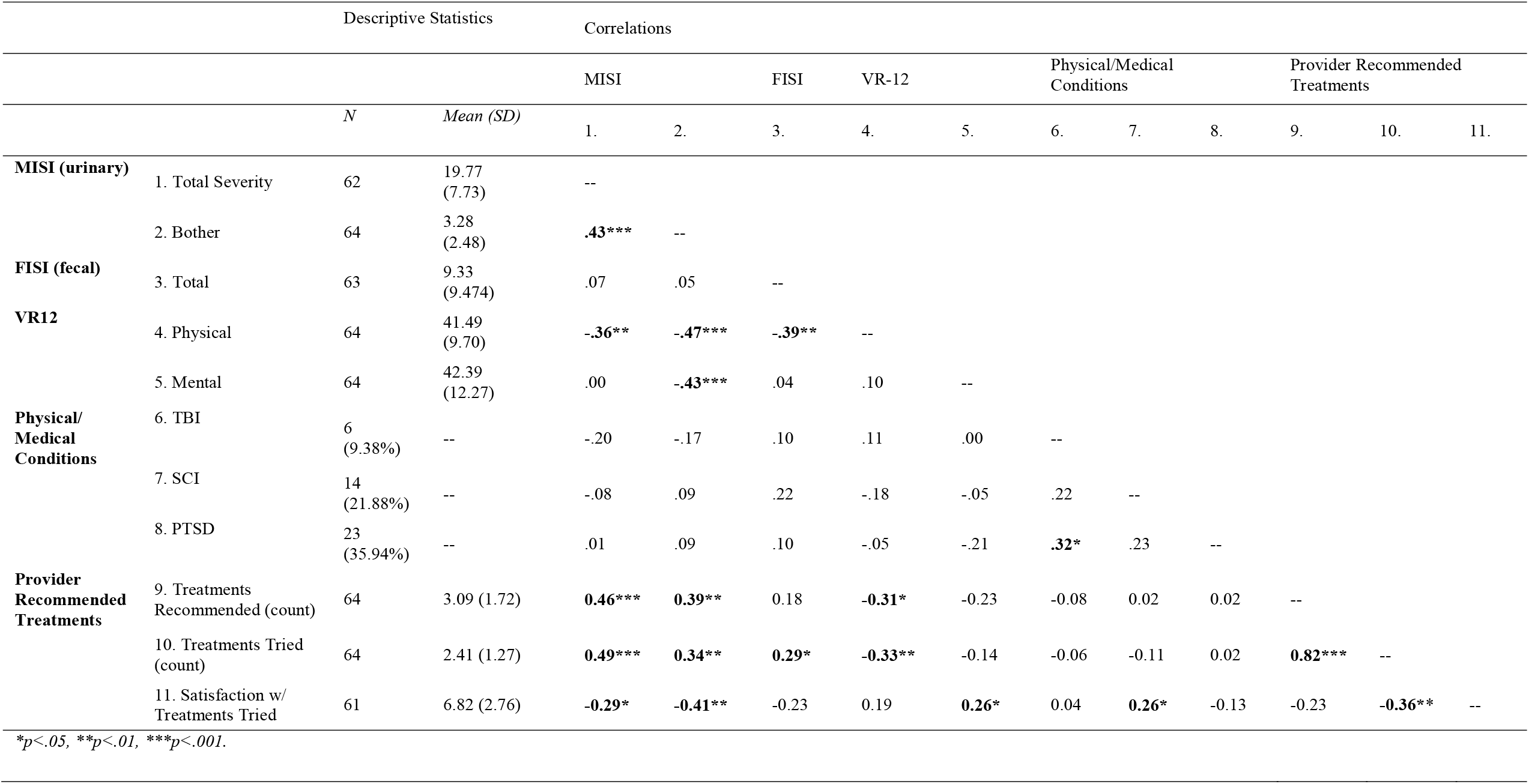
Zero-Order Correlations for Veteran Measures (*n* = 64)

Indeed, the VR-12 physical domain correlated significantly and negatively with urinary incontinence severity (*r* = −.36, *p <* .01), urinary incontinence bother (*r* = −.47, *p <* .001), and fecal incontinence severity (*r* = −.39, < .01). The VR-12 physical domain also correlated significantly and negatively with the number of treatments recommended by providers (*r* = −.31, *p <* .05) and number of treatments tried (*r* = −.33, *p <* .01). Additionally, mental quality of life correlated negatively with urinary incontinence bother (*r* = −.43, *p* < .001). Overall, findings indicate that Veteran quality of life is negatively related to symptoms of incontinence.

### Caregiver Burden

Caregiver burden correlated positively with caregiver perception of urinary incontinence bother (*r* = .46, *p <* .01); the greater the level of caregiver burden, the more the caregiver perceived that the Veteran is bothered by their urinary incontinence. Caregiver burden also correlated significantly and negatively with satisfaction with treatments tried (*r* = −.38, *p <* .05); the greater the caregiver’s burden, the lower their satisfaction with the treatments tried. See Table 3 in the Supplementary Material for more information.

### Veteran and Caregiver Dyads: Perception of Incontinence

Table 5 presents zero-order correlations between Veteran- and caregiver-reported variables within the Veteran-caregiver dyad subsample. Due to the low sample size of the dyads (*n* = 13), the strength of the correlations should be interpreted with caution. Veteran’s perceptions of UI severity (i.e., measured by MISI) correlated positively with caregiver perceptions of UI severity; the greater the Veterans’ urinary incontinence severity, the higher the caregivers’ perception of urinary incontinence severity. Caregiver perception of urinary incontinence bother correlated negatively with Veteran mental quality of life; the higher the Veteran’s mental quality of life, the lower the caregiver’s perception of how bothersome the Veteran’s urinary incontinence is.

**Table 5.**
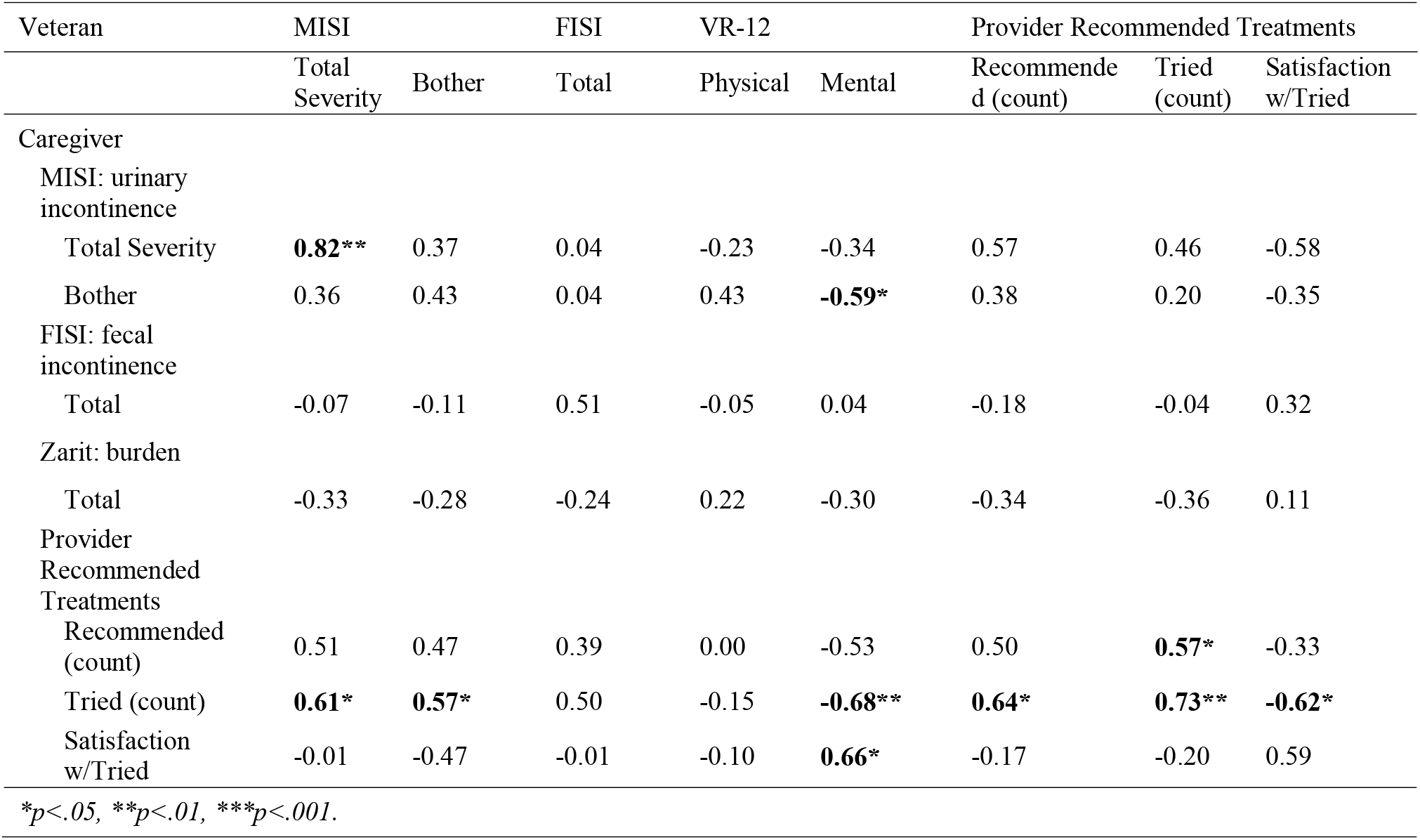
Zero-Order Correlations for Veteran & Caregiver Measures (*n* =13 dyads)

| Veteran | MISI |  | FISI | VR-12 |  | Provider Recommended Treatments |  |  |
| --- | --- | --- | --- | --- | --- | --- | --- | --- |
|  | Total Severity | Bother | Total | Physical | Mental | Recommended (count) | Tried (count) | Satisfaction w/Tried |
| Caregiver |  |  |  |  |  |  |  |  |
| MISI: urinary incontinence |  |  |  |  |  |  |  |  |
| Total Severity | **0.82\*\*** | 0.37 | 0.04 | -0.23 | -0.34 | 0.57 | 0.46 | -0.58 |
| Bother | 0.36 | 0.43 | 0.04 | 0.43 | **-0.59\*** | 0.38 | 0.20 | -0.35 |
| FISI: fecal incontinence |  |  |  |  |  |  |  |  |
| Total | -0.07 | -0.11 | 0.51 | -0.05 | 0.04 | -0.18 | -0.04 | 0.32 |
| Zarit: burden |  |  |  |  |  |  |  |  |
| Total | -0.33 | -0.28 | -0.24 | 0.22 | -0.30 | -0.34 | -0.36 | 0.11 |
| Provider Recommended Treatments |  |  |  |  |  |  |  |  |
| Recommended (count) | 0.51 | 0.47 | 0.39 | 0.00 | -0.53 | 0.50 | **0.57\*** | -0.33 |
| Tried (count) | **0.61\*** | **0.57\*** | 0.50 | -0.15 | **-0.68\*\*** | **0.64\*** | **0.73\*\*** | **-0.62\*** |
| Satisfaction w/Tried | -0.01 | -0.47 | -0.01 | -0.10 | **0.66\*** | -0.17 | -0.20 | 0.59 |

\**p*<.05, \*\**p*<.01, \*\*\**p*<.001.

While in the caregiver subsample, burden correlated significantly with caregiver-reported perception of Veteran urinary incontinence bother, in the dyad subsample, there was no evidence of significant relationships between caregiver burden and any Veteran-reported measures of incontinence, nor for quality of life.

### Veteran and Caregiver Dyads: Treatments & Satisfaction

Of the correlations between Veteran-reported and caregiver-reported treatments and satisfaction, the strongest correlation was between Veteran-reported and caregiver-reported number of treatments tried (*r* = .73, *p* < .01); the greater the number of recommendations the Veteran reported, the greater the number of recommendations the caregiver reported.

### Qualitative results

Overall, the quantitative data demonstrated that incontinence symptoms significantly and negatively correlated with Veteran’s quality of life. The qualitative data expands upon these findings by highlighting how the participants perceived and managed their incontinence, especially regarding their treatment experiences. In 26 interviews with 18 Veterans and 8 caregivers, qualitative data revealed 6 primary themes related to Veteran and caregiver incontinence experiences: 1) Incontinence knowledge 2) Seeking care for incontinence 3) Satisfaction with healthcare providers 4) Satisfaction with treatments 5) Satisfaction with products and 6) Quality of life.

#### Theme 1: Veteran and caregiver knowledge about incontinence

Participants were queried about their knowledge about incontinence prior to their onset of incontinence and their reactions to their symptom development. Most (63%) heard little or nothing about incontinence before their illness onset. One male Veteran explained, “Well I didn’t know it [incontinence] existed. I just thought I was late getting up to go to the bathroom.” Those that had heard something about incontinence said that they believed it was an inevitability when you get older, are a woman, give birth, or have a hysterectomy, and only later learned that incontinence was a separate medical condition. A female Veteran explained this:

> “I had a total hysterectomy…I couldn’t hold my urine as I did before my procedure. I thought it was my body trying to repair itself and heal, but it got worse over time…When I went to see my doctor to return to work, I asked him was it normal for me to pee on myself. He said no, it wasn’t normal.”
>
> Like both Veterans quoted above, 26% of the Veterans interviewed did not recognize their symptoms as incontinence at the onset of symptoms. Almost half (43%) directly associated the incidence of incontinence with another medical condition or surgical procedure.

#### Theme 2: Seeking care for incontinence symptoms

When some participants first noticed there was an issue with bladder or bowel control, 32% reported that they saw a doctor. An equal number bought products on their own, decided to wait and see it if happened again, or noted that they did not tell anyone that it happened. Some participants never sought medical assistance, even after experiencing the issue for decades. Regardless of initial hesitation to seek treatment, most participants (80%) reported not feeling uncomfortable speaking about their incontinence to any of their medical providers. Participants who did feel uncomfortable had specific providers that they did not want to speak with. When asked what medical assistance participants had sought for their incontinence symptoms, 65% said that they saw their primary care provider first. Similarly, 74% of participants spoke to their primary care provider about the issue first before any other medical professional, friend, or family member. Most of the caregivers learned about their Veteran’s symptoms because of the need to do additional laundry or other cleaning, not because the Veteran told them about the issue.

#### Theme 3: Satisfaction with healthcare providers

Over 55% of participants reported positive experiences discussing the issue of incontinence with a healthcare provider. Commenting on his experience discussing incontinence with his primary care provider, one male Veteran expressed, “I fully understood what she was talking about. She explains something and then waits to see if you understand. She is very straightforward about it. Very attentive provider.” Many participants (41%) were satisfied with the amount of time and attention they received from their provider, including one female caregiver who said, “Overall, they [VA] do a wonderful job. I can just call them and tell them what the problem is and they get right on it. They don’t stop for nothing.”

An additional 36% of participants explained that they experienced explicitly negative experiences when discussing their incontinence with a healthcare provider. One male Veteran stated, “Seems like in the military they don’t take it seriously. They just basically tell you to deal with it.” Other reasons for negative experiences included insufficient discussion of treatment alternatives, or that the provider was “not helpful” or “not listening.” Several participants stated that their healthcare provider did not provide enough time to discuss the issue or did not give enough attention to the issue. Some participants felt negatively about the explanations given to them by healthcare providers on how to manage or reduce incontinence. A female caregiver explained her concern when she said, “Medically speaking I think he handled it the way that he should handle it, you know what I’m saying, because he offered medicine. But sometimes there’s more alternatives.” This caregiver also said she felt like the provider wanted to write a prescription for the issue to get her and her Veteran out of the office, instead of discussing other treatments.

When asked how communication with healthcare providers could be improved, participants suggested that that providers needed a better understanding of all of the treatments and products for incontinence (29%), that providers need to communicate more clearly with Veterans (21%), that doctors need a better understanding of female incontinence (14%), and that doctors need to listen more to their Veteran patients (14%). Some participants were unsure how to improve communication with their provider, because they had been avoiding the conversation about their incontinence for so long. For example, one male Veteran said, “I don’t know how it could be improved. I know that nobody asked me about it. Maybe they should inquire about it as part of their list of questions. I had to bring up the whole thing when I was finally at my wits end as to what to do about it. I’ve been dealing on my own for decades.”

#### Theme 4: Satisfaction with treatments for incontinence

When asked about difficulties Veterans had experienced in following treatment recommendations from healthcare providers, only 21% of participants said that they had no issues following the recommendations of their healthcare provider, while 33% perceived being given recommendations that they did not think were appropriate for their them or their symptoms. A female caregiver for a male Veteran with incontinence explained her experience:

> “Typically, for brain injury their go-to is to just use a condom catheter for anybody and everybody. And that isn’t really an option for us because, well, [the Veteran] is mobile and…is cognitive enough to know that something is attached to him and he pulls it off.”

Some Veterans stated they were not given enough supplies to manage their symptoms. One male Veteran asked, “How in the world does the pharmacy expect me to use two pads per day when you want me to catheterize 5-6 times a day? What am I supposed to do, put on a soiled pad just after getting myself all cleaned up?”

Other participants were dissatisfied with their treatments for incontinence because they were given recommendations that they felt were too embarrassing to follow, including medication that still resulted in periodic bladder incontinence, using menstrual pads (for men), and using products that other people could notice. A male Veteran described his discomfort in saying, “[Diapers are] bulky and uncomfortable…I thought, ‘I ain’t going this route, no’.” Some participants felt the recommendations they received were too inconvenient to follow, including catheters that were the wrong size and having to visit the provider multiple times a year to get supplies. Finally, some participants were so frustrated with the process of seeking treatment that they never received appropriate recommendations, including one woman who was repeatedly referred to a urologist by her primary care provider, and then referred back to the primary care provider by the urologist. Even though only 21% received a treatment plan that they were satisfied with, almost half of participants (44%) reported not deviating from their provider’s recommended treatment plan.

#### Theme 5: Satisfaction with incontinence products

Most participants (79%) described that they learned about the products they were using from their healthcare provider and many (22%) said that they will use whatever product the VA sends them, regardless of brand. When asked what products they used, some participants were using pads that fit in their underwear (32%), using incontinence briefs (20%), and using catheters (20%). Reasons for using different products varied and were related to the lifestyle of the participant or the Veteran they care for. For example, one female caregiver expressed, “I would say using diapers [incontinence briefs] has definitely increased quality of life. It’s easier, you know, than the alternative which for us was the condom cath. So, for us it increases quality of life.” Alternately, a female Veteran explained that, from her perspective, “Oh no, it’s not diaper time yet. No. I’d wear three pads before I’d wear a diaper. I have a lot of friends who have incontinence and they wear those briefs, those throw away panties and stuff like that. I can’t do that. To me, I’m not ready for that.” A few participants did not use any products to manage their symptoms, such as the male Veteran who said, “I’m not using anything yet. I don’t want to really, if you want to know the truth. It makes me feel like an old man.”

Participants had many ideas about how to improve products. For example, 39% said that they would like products to be less bulky. Similarly, 39% recommended products to be available in more sizes, and 11% would like products to be more masculine. Other product improvement suggestions included that briefs be less saggy, absorb liquid better, or control odor better. Several female Veterans described their struggle with fitting products under their clothing, such as the Veteran who said, “I try to use the pads, like when you’re on your period? I try to use those when I go out, because…to me I feel like everybody can see what I’m wearing…whether I wear the diaper or the pads, I still wear panties over them. So, I try to not show the diaper.” Other product preferences were related to the utility of the product, which was expressed by a female caregiver who explained that she will avoid products that “promote skin breakdown, and do not hold urine effectively for an adult, or someone who is truly 100% incontinent.” She will seek products that “feel more like real underwear” because “they’re more comfortable, they’re softer on the body, and they have more of a feel-tight design to them.”

Most participants (71%) said that they were getting all of their supplies from the VA, and therefore did not feel an economic impact on their family. However, several mentioned that if they were not getting products from the VA, they would not be able to afford them. An additional 28% of participants only bought incontinence supplies periodically or did not buy them altogether because they could not afford to. Some of these participants did not know they could get incontinence supplies through the VA. Several participants were purchasing products in addition to what was supplied to them by the VA.

#### Theme 6: Quality of life

When prompted to discuss their experiences with incontinence, many participants stressed that there is an emotional toll to experiencing incontinence that must be taken into consideration. One male Veteran explained that he is “always on edge” any time he leaves his home, and that it’s “not worth living like that.” One female Veteran expressed that she saw her incontinence as a threat to her marriage, as she had been hiding it from her husband for about a decade. Another female Veteran described the way her life had changed following the onset of her incontinence symptoms when she said:

> “This incontinence thing? I’ve got to tell you, mentally, it’s breaking me down. I’m actually seeing a psychiatrist right now because this has just tore my life apart. I never thought I would be some 69-year-old woman, laying in a bed, messing her pants. That’s not how I saw myself. I was always active, I was out in the community, making friends, going places, doing things. And now I don’t go anywhere or do anything, I’m just like a lump.”

These participants emphasized that they were highly concerned with other people’s perceptions of their incontinence and were concerned the incontinence could be discovered because of odor or because the products they use to manage their symptoms were visible to others.

## Discussion

This study sought to determine the feasibility of recruitment strategies, methodology, and workflow processes to study Veteran incontinence. Using a telephone survey methodology over the course of 1-year, this study collected data from 100 participants, including 64 Veteran patients and 36 caregivers. This study demonstrated that there is strong interest in the topic from the sample population. Surprisingly, there was much stronger interest in the topic of incontinence than we anticipated, with both Veterans and caregivers being eager to talk with us about their experiences. Out of 123 participants who met eligibility criteria, 114 (92%) agreed to participate in the study. Further, 74 participants from the final sample of 100 indicated they would be interested in participating in future studies on incontinence. Overall, this study demonstrates that recruiting a substantial sample of Veterans with incontinence, and their caregivers, is feasible using our recruitment methods and strategies. This will be used to inform a larger future study on this topic.

This study also aimed to provide preliminary data about incontinence experiences among Veterans and their caregivers, using quantitative and qualitative data. Results illustrate that incontinence must be recognized as a significant quality of life issue for Veterans and their caregivers. In our sample, Veterans reported a reduced quality of life compared to the general population (Kazis et al., 2006). This finding was true both for physical quality of life, which was correlated to incontinence severity, and mental quality of life, which similarly was correlated to the bother caused by incontinence. The qualitative data converged with the quantitative data about these quality of life detriments. Veterans openly described incontinence as limiting their social and employment opportunities, even forcing some participants to remain housebound while they were experiencing symptoms (for more discussion on qualitative findings, see XXXX, 2020). Caregivers generally provided similar assessments as Veterans in the quantitative findings, indicated by significant correlations between Veteran and caregiver perceptions of UI severity, bother, and quality of life. Although more research is needed with larger samples and additional means to establish agreement, this preliminary finding suggests that caregivers man potentially provide relatively reliable symptom information in agreement with Veterans’ symptom experiences.

Results from the qualitative findings highlight that incontinence often represents a difficult topic for Veterans to discuss with their caregivers or family members, and Veterans may conceal or underreport their symptoms from them (Smoger et al., 2000; Devendorf et al., 2020). From the qualitative findings, caregivers commonly discovered that their Veteran was experiencing incontinence because they saw evidence of the symptoms when the Veteran had an accident, or while doing laundry. None of the caregivers said that they learned about the Veteran’s incontinence through a discussion with their Veteran. Several Veterans without caregivers mentioned hiding evidence from their children or spouses. Reasons for not disclosing included embarrassment, shame, and lack of knowledge that incontinence was a serious medical issue (Devendorf et al., 2020). Moreover, many participants expressed that they did not recognize their symptoms as incontinence when they first occurred.

Relatedly, the quantitative and qualitative results revealed that many Veterans delay seeking incontinence treatment for long periods of time. In the total Veteran sample, 28% experienced incontinence symptoms for 1-5 years, and 67% experienced symptoms for 5-years of more. However, 44% of Veterans waited at least 1-year or more to discuss incontinence with a VA provider. These findings align with findings in civilian samples with incontinence (Koch, 2006; Taylor, Cahill, & Rizk, 2014). Fortunately, almost all Veterans in our sample eventually sought treatment, and for those Veterans who initiated treatment at the VA, 55% stated that they had positive experiences, and 80% said that there was no medical provider that they felt uncomfortable speaking with about incontinence. These findings highlight the need for more general education among Veterans and caregivers about what incontinence is, when it may occur, and what to do if it happens. Such educational interventions could occur in healthcare settings after procedures that generate elevated risk for incontinence, including procedures for hysterectomies.

In fact, the importance of having providers that are well informed about incontinence and its treatment is underscored by the high percentage of Veterans who sought incontinence treatment; 92% of our Veteran sample reported ever speaking with a VA provider about incontinence while only 42% reported speaking with a non-VA provider. In the VA, primary care providers could serve as Veterans’ first introduction to education about incontinence and associated products. Discussions with these providers could prepare Veterans and their caregivers to recognize incontinence symptoms, normalize discussion of the topic, and improve treatment-seeking behaviors. Provider education may also improve patient-provider relationships (Helewa et al., 2017; Taylor & Cahill, 2018). The qualitative data suggest that, while Veterans reported feeling comfortable talking with their provider, Veterans also commonly desired better communication with their providers about incontinence, including obtaining more appropriate and tailored treatment options.

In the total sample, 97% of Veterans who sought treatment from a medical provider tried their providers recommendations. Veterans mostly tried over-the-counter products (86%), medications (61%), physical therapy or saw a specialist for therapy (34%), protective creams or ointments (30%), surgery (13%), among other strategies (see supplementary material for details). These results demonstrate that providers provide an array of different treatment options. Similarly, that Veterans are willing to try these treatments may signify considerable symptom severity, and trust in their provider. This is evinced by the quantitative results that show that higher incontinence severity and bother, and lower quality of life, are associated with a greater number of recommendations from the provider (as reported by Veterans). Additionally, the number of Veteran-reported treatments tried correlates strongly and positively with the number of Veteran-reported treatments recommended. Overall, this study suggests that Veterans are willing to follow recommendations from providers, and providers are responding to the Veterans’ severity of symptoms and quality of life in making their recommendations.

### Limitations and Strengths

In considering this study’s findings, it is important to note its limitations. First, because this was a pilot study that aimed to assess the feasibility of our recruitment procedures for this sample, our study was unable to recruit a large, representative sample of Veterans. Second, the cross-sectional design precludes our ability to understand causal relationships across variables. Third, our sample size for the quantitative portion was relatively small for caregivers and moderate for Veterans, limiting our power to detect reliable relationships across variables. Relatedly, our sample size of 26 participants was moderate for our qualitative portion, and we were unable to recruit male caregivers of Veterans with incontinence. Additionally, we were unable to determine whether any relationships between caregiver burden and Veteran-reported (not caregiver-perceived) symptom severity, bother, or quality of life exists, or whether there was simply insufficient data to detect them.

Despite these limitations, our study has notable strengths. To our knowledge, this study was the first to document Veterans’ management strategies, quality of life, and treatment experiences in managing incontinence using a mixed-methods approach. Our study fills a needed research gap through including Veteran and caregivers’ accounts of incontinence. That our study includes participants with UI and FI increases our ability to generalize our findings across incontinence. Additionally, our qualitative interviews provided us with rich, in-depth information that supplemented our quantitative surveys. Given the sensitivity and stigma surrounding incontinence (Devendorf et al., 2020), using qualitative interviews to supplement quantitative data will be important for the field moving forward. Lastly, our study provides a groundwork for conducting a larger study moving forward.

An important strength of this study was the very active Veteran Engagement Council (VEC) at DEIDENTIFIED FOR REVIEW (the local study site). This group meets once a month for knowledge sharing between the Veteran population the VA serves (including their caregivers) and research or project planning teams. VA researchers can come talk to the VEC about a proposal idea being developed and gain valuable insight to best approaches, priority areas of interest, and respectful plans for recruitment and participation. The feasibility project team met with the VEC on 3 occasions: 1. Prior to research study submission for regulatory approvals (during project proposal and phone survey/interview guide development); 2. After regulatory approvals and revisions, when the final survey and interview guides were completed (to ask for further feedback and suggestions for recruitment strategies); 3. After the feasibility study had been mostly completed – to review preliminary survey and interview results (and the poster we presented at the local rehabilitation fair) to thank them for their interest and feedback. The involvement of the VEC as a resource group related to the local research center was invaluable. Having Veteran and caregiver input helped in tweaking the wording of phone survey questions, recruitment letters, and interview questions, and asking questions the VEC felt were most pertinent for the research topic.

### Conclusion and Future Directions

This study demonstrates that incontinence among Veterans is associated with significant impairment across physical, mental, and social domains. Veterans in this study generally concealed their incontinence from friends, family, and providers, although most reported having positive VA healthcare experiences once they sought treatment. Key next steps involve conducting studies that are larger, more representative, and should include longitudinal studies following Veterans with incontinence, and their caregivers, over time. It will be important to understand the processes (e.g., psychological, behavioral, environmental) that help Veterans cope with accepting and managing their incontinence over time. Likewise, it will be essential for clinicians to build a rapport with Veterans, encouraging earlier disclosure of incontinence symptoms.

Our study suggests that intervening at the healthcare level by educating providers and systematizing inquiry into potential incontinence for higher risk populations would be a fruitful avenue to explore. Especially at the VA, providers can serve as gatekeepers for health and referral information for Veterans. In addition to further background knowledge about incontinence, healthcare providers, Veterans, and caregivers all need greater knowledge about possible treatment avenues, including available products and for whom certain products are appropriate. Many participants had specific complaints about treatments or products they were using or desires for greater product availability without realizing that options already exist to fulfill their needs. This was true for products like pads and briefs, as well as for catheters. Future studies could explore how to mitigate the chance that a provider delivers unhelpful or inappropriate information. Additionally, future studies could explore how to educate Veterans about incontinence through options that may not require having a conversation with a human (to minimize embarrassment) such as e-health and automated screening options on VA websites. *This will be especially important when considering that individuals* with incontinence experience self-stigmatizing attitudes like shame, embarrassment, and fear (Devendorf et al., 2020).

Ultimately, incontinence among Veterans and their caregivers is a severely understudied topic. To provide effective treatments and intervention for Veterans, it will be important to better understand the lived experiences associated with this group. We hope this first investigation will motivate additional research in this area.

## Data Availability

Additional information about data and analysis may be provided upon request to the first author.

## Acknowledgements and disclosures

This material is the result of work supported with resources and the use of facilities at the James A. Haley Veterans’ Hospital. The contents do not represent the views of the Department of Veterans Affairs or the United States Government. This research was funded by a research grant from DOMTAR, Inc.

This material is the result of work supported with resources and the use of facilities at the DEIDENTIFIED FOR REVIEW PURPOSES. The contents do not represent the views of DEIDENTIFIED FOR REVIEW PURPOSES. This research was funded by a research grant from DEIDENTIFIED FOR REVIEW PURPOSES.

## Supplementary Material

**Supplement Table 1.**
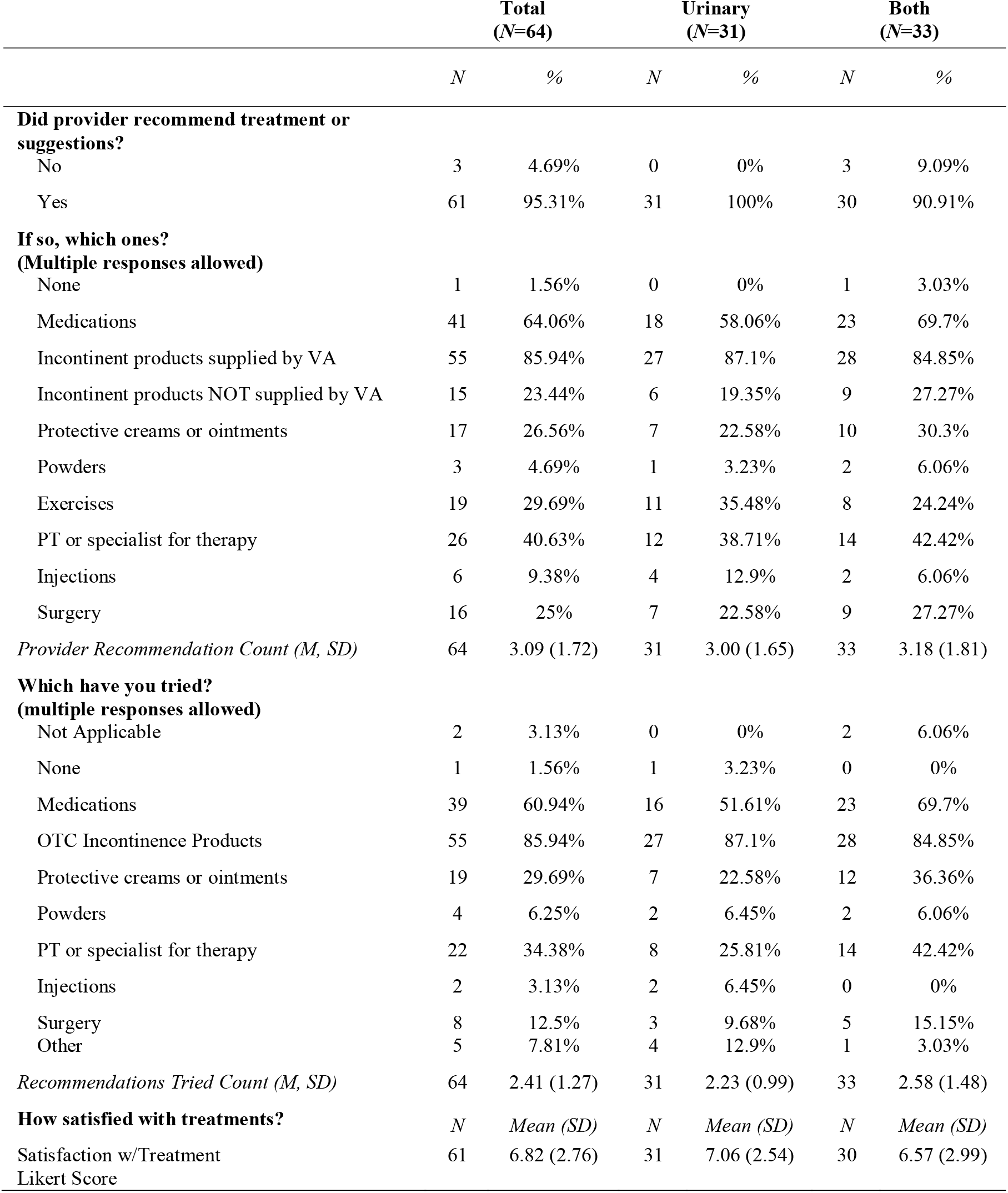
Healthcare Provider Recommendations and Satisfaction (as reported by Veterans) by Incontinence Type.

|  | Total<br>(N=64) |  | Urinary<br>(N=31) |  | Both<br>(N=33) |  |
| --- | --- | --- | --- | --- | --- | --- |
|  | N | % | N | % | N | % |
| **Did provider recommend treatment or suggestions?** |  |  |  |  |  |  |
| No | 3 | 4.69% | 0 | 0% | 3 | 9.09% |
| Yes | 61 | 95.31% | 31 | 100% | 30 | 90.91% |
| **If so, which ones?**<br>**(Multiple responses allowed)** |  |  |  |  |  |  |
| None | 1 | 1.56% | 0 | 0% | 1 | 3.03% |
| Medications | 41 | 64.06% | 18 | 58.06% | 23 | 69.7% |
| Incontinent products supplied by VA | 55 | 85.94% | 27 | 87.1% | 28 | 84.85% |
| Incontinent products NOT supplied by VA | 15 | 23.44% | 6 | 19.35% | 9 | 27.27% |
| Protective creams or ointments | 17 | 26.56% | 7 | 22.58% | 10 | 30.3% |
| Powders | 3 | 4.69% | 1 | 3.23% | 2 | 6.06% |
| Exercises | 19 | 29.69% | 11 | 35.48% | 8 | 24.24% |
| PT or specialist for therapy | 26 | 40.63% | 12 | 38.71% | 14 | 42.42% |
| Injections | 6 | 9.38% | 4 | 12.9% | 2 | 6.06% |
| Surgery | 16 | 25% | 7 | 22.58% | 9 | 27.27% |
| *Provider Recommendation Count (M, SD)* | 64 | 3.09 (1.72) | 31 | 3.00 (1.65) | 33 | 3.18 (1.81) |
| **Which have you tried?**<br>**(multiple responses allowed)** |  |  |  |  |  |  |
| Not Applicable | 2 | 3.13% | 0 | 0% | 2 | 6.06% |
| None | 1 | 1.56% | 1 | 3.23% | 0 | 0% |
| Medications | 39 | 60.94% | 16 | 51.61% | 23 | 69.7% |
| OTC Incontinence Products | 55 | 85.94% | 27 | 87.1% | 28 | 84.85% |
| Protective creams or ointments | 19 | 29.69% | 7 | 22.58% | 12 | 36.36% |
| Powders | 4 | 6.25% | 2 | 6.45% | 2 | 6.06% |
| PT or specialist for therapy | 22 | 34.38% | 8 | 25.81% | 14 | 42.42% |
| Injections | 2 | 3.13% | 2 | 6.45% | 0 | 0% |
| Surgery | 8 | 12.5% | 3 | 9.68% | 5 | 15.15% |
| Other | 5 | 7.81% | 4 | 12.9% | 1 | 3.03% |
| *Recommendations Tried Count (M, SD)* | 64 | 2.41 (1.27) | 31 | 2.23 (0.99) | 33 | 2.58 (1.48) |
| **How satisfied with treatments?** | N | Mean (SD) | N | Mean (SD) | N | Mean (SD) |
| Satisfaction w/Treatment Likert Score | 61 | 6.82 (2.76) | 31 | 7.06 (2.54) | 30 | 6.57 (2.99) |

**Supplement Table 2.**
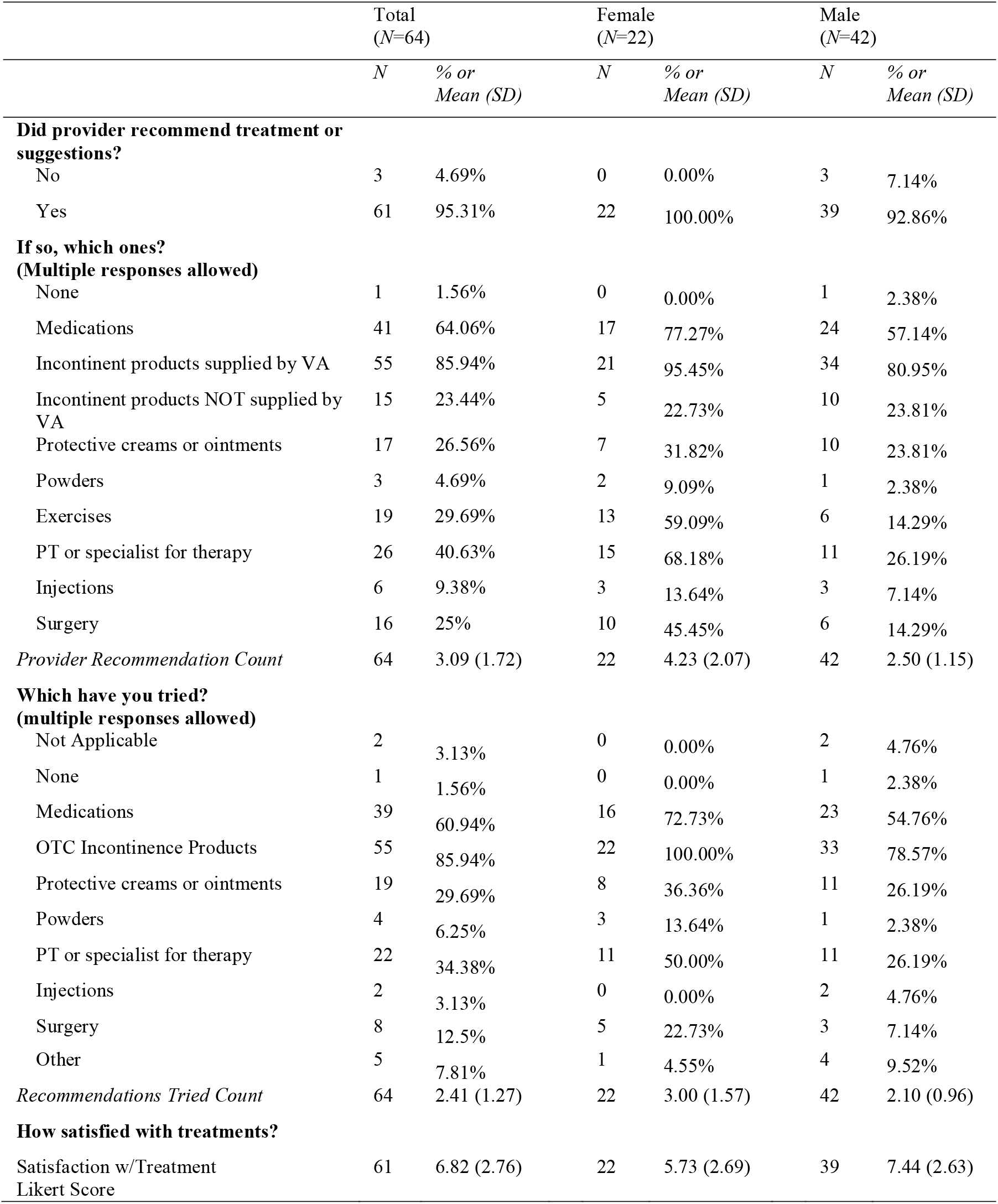
Healthcare Provider Recommendations (as reported by Veterans) by Veteran Gender.

**Table 3.**
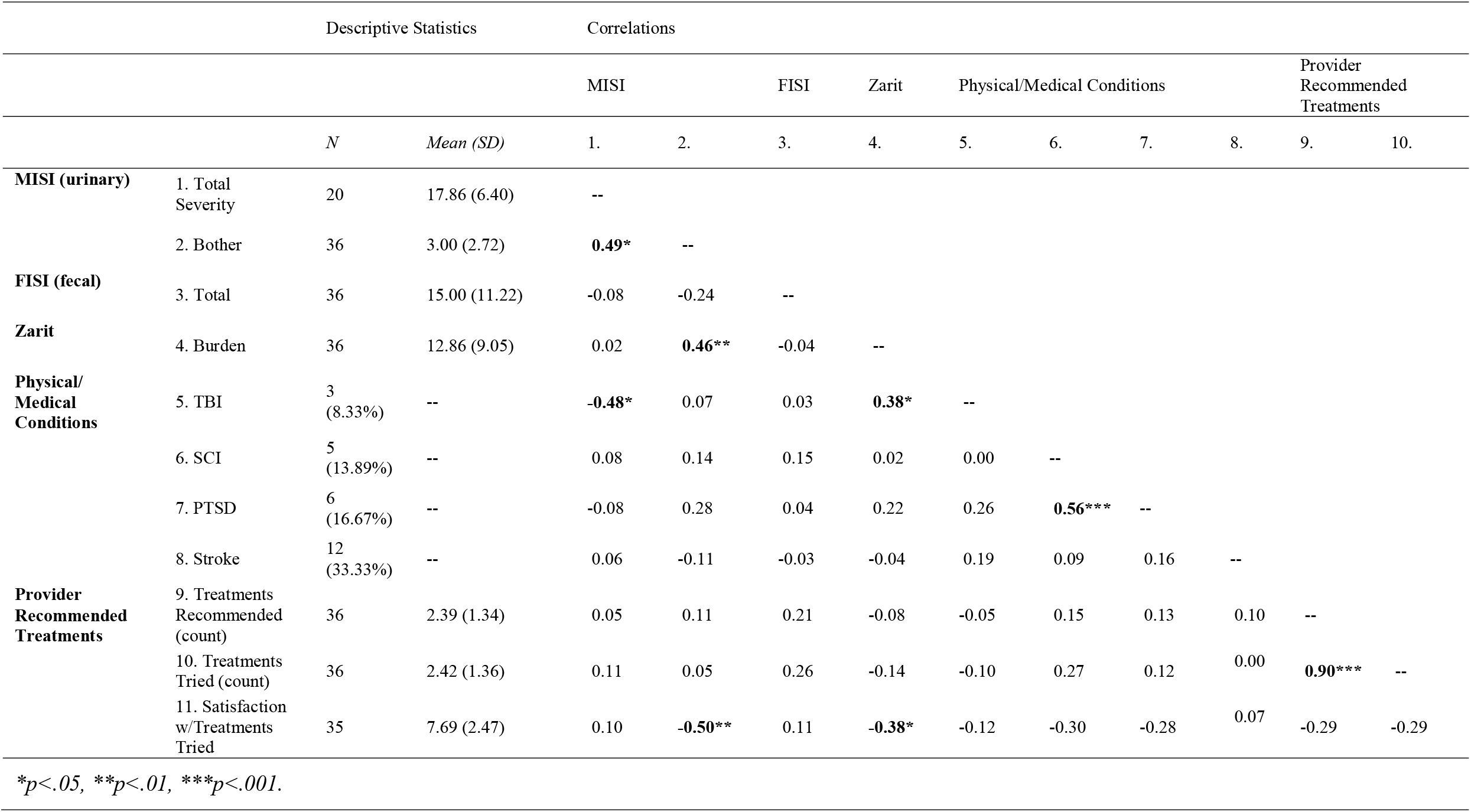
Zero-Order Correlations for Caregiver Measures (n = 36)

